# Decrease in multiple complement protein levels is associated with the development of islet autoimmunity and type 1 diabetes

**DOI:** 10.1101/2023.07.13.23292628

**Authors:** Bobbie-Jo M Webb-Robertson, Ernesto S Nakayasu, Fran Dong, Kathy C Waugh, Javier Flores, Lisa M Bramer, Athena Schepmoes, Yuqian Gao, Thomas Fillmore, Suna Onengut-Gumuscu, Ashley Frazer-Abel, Stephen S Rich, V. Michael Holers, Thomas O Metz, Marian J Rewers

## Abstract

Type 1 diabetes (T1D) is a chronic condition caused by autoimmune destruction of the insulin-producing pancreatic β-cells. While it is known that gene-environment interactions play a key role in triggering the autoimmune process leading to T1D, the pathogenic mechanism leading to the appearance of islet autoantibodies - biomarkers of autoimmunity – is poorly understood. Here we show that disruption of the complement system precedes the detection of islet autoantibodies and persists through disease onset. Our results suggest that children who exhibit islet autoimmunity and progress to clinical T1D have lower complement protein levels relative to those who do not progress within a similar timeframe. Thus, the complement pathway, an understudied mechanistic and therapeutic target in T1D, merits increased attention for use as protein biomarkers of prediction and potentially prevention of T1D.

## Introduction

Clinically onset of type 1 diabetes (T1D) is preceded by a period of islet autoimmunity (IA) that is marked by the appearance of circulating autoantibodies against islet autoantigens^1–4^. While there is a consensus that chronic β-cell autoimmune destruction is triggered by interactions of genetic, genomic, and environmental factors, the pathogenesis of the initiation and progression of the disease is still largely unknown. Identification of biomarkers that predict triggering of IA in at-risk individuals, as well as progression from IA to clinical diabetes, may give clues into the etiology of this complex disease. Dysregulation of the complement system can affect innate immunity and has been noted in macular degeneration^5^, cancer^6^, and autoimmune disorders, such as systemic lupus erythematosus^7, 8^. Several non-hypothesis driven discovery studies have suggested that the complement system may play a role in the etiology of T1D^9–13^. For example, local production of complement component C3 is an important survival mechanism in β cells under a proinflammatory assault. In response to interleukin-1β and interferon-γ, C3 expression increases in rodent and human β cells^14^. This increased C3 expression may enhance autophagy - a protective response to β-cell stress - and improve β-cell function^15^.

Proteomics is a core discovery technology utilized to identify biomarkers of disease, as well as gain insight into the molecular processes driving disease progression, that has been employed previously to study the pathogenesis of T1D^9, 11, 12, 16, 17^. These studies have focused on a combination of global and targeted analyses in cohorts evaluating time-based measurements in the progression of T1D. Changes in levels of multiple complement proteins, such as C4, C3, C2 and C1r, have been reported^9, 11^, although the directional changes varied and thus a cohesive pattern associated with the complement system and T1D is has not yet emerged.

Here we present an investigation of the relationship specifically between the complement system and progression to IA and T1D. To accomplish this task, we analyzed complement proteins in a long-standing birth cohort of high-risk children, the Diabetes Autoimmunity Study in the Young (DAISY). DAISY defines high-risk status as having a first-degree relative with T1D or high genetic risk HLA (Human Leukocyte Antigen) genotype^18, 19^. Complement proteins were measured as their constituent peptides using selected reaction monitoring (SRM)-based targeted proteomics^20^. In addition, multiple complement proteins, of which approximately half overlap with the SRM measured proteins, were also sent for quantitative complement testing using commercial immunoassays in a CAP/CLIA accredited laboratory^21^ (Exsera BioLabs), referred herein as Exsera. Both approaches are well established methods for quantitative proteomics and offer confirmatory evidence.

## Results

A total of 172 children from the DAISY study with multiple plasma samples collected over time, with up to 23 years of follow-up, were characterized via proteomics analysis, **Fig 1**. Of the children there were 40 controls (**Fig. 1A**) and 132 cases (**Fig. 1B-D**). All 132 cases had measurements across time relative to IA. Sampling was not consistent for all children. There were 47 of the children who had samples taken and evaluated prior to IA (Pre-IA), represented as p-xx (**Fig. 1B**), and 131 children had measurements at or after IA, but prior to diagnosis of clinical T1D (Post-IA), represented as i-xxx (**Fig. 1B-D**). The control children were frequency matched on HLA genotypes and age and sex (**Table 1**) with an observed lower frequency of first degree relatives within the control group versus the cases. The Pre- and Post-IA proteomics measurements highlighted in **Fig. 1B-D** were compared to the control samples in **Fig 1A** using a linear mixed model to evaluate association of complement proteins with these two important events in the progression of T1D.

**Fig. 1:**
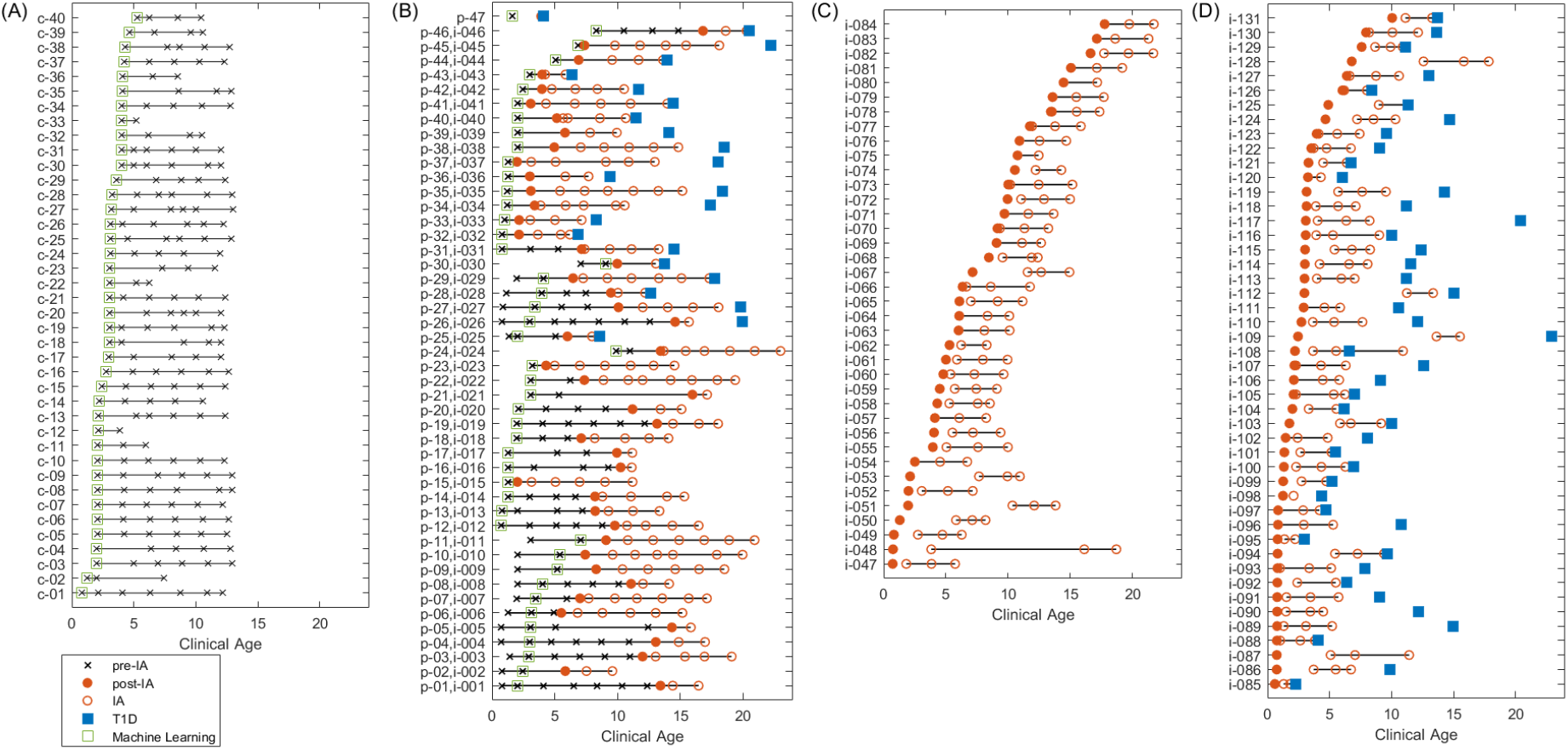
Clinical age that samples were analyzed. For each child associated with the study key endpoints highlighted, including if the sample is prior to (x) or after (o) IA, the point of IA (●), the age of T1D diagnosis (▪), and the single sample used for the machine learning (ML) analysis (□). Plot is separated into (A) 40 control samples, (B) 47 pre-IA samples, of which 24 were not diagnosed with T1D during the study and 23 were, and 46 of these 47 also had post-IA measurements, (C) 38 post-IA samples who were not diagnosed with T1D during the study, and (D) 47 post-IA samples who were diagnosed with T1D during the study.

**Table 1:**
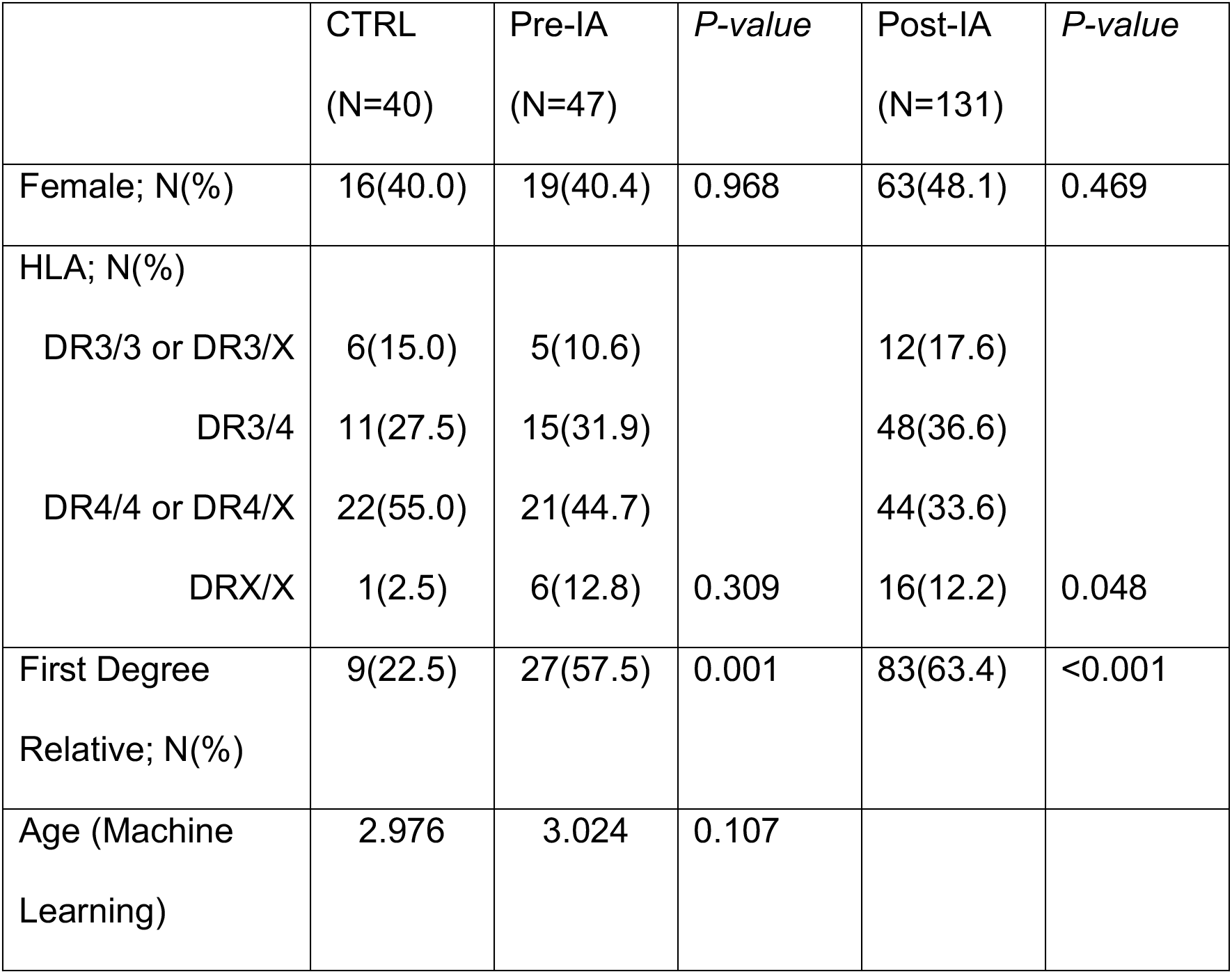
Demographics of the DAISY cohort for the Control, Pre-IA, Post-IA and Pre-T1D samples. The Pre-IA, Post-IA, and Pre-T1D are compared to control where sex, HLA and First Degree Relative are compared to the control group via ξ2 test of independence and age is compared via a two-sample-test.

### Association of SRM Complement Proteins with IA and T1D

The SRM proteomics experiment measured 19 complement proteins, most of which were identified by at least two unique peptides. Protein level data is presented as the average of the measured peptides. The statistical results are presented visually as an average log2 fold-change at five different age ranges across all subjects. The association of the relationship of IA relative to control children shows a distinct decrease for most complement proteins, **Fig. 2**. The overall pattern of a decrease in complement proteins is persistent both before and after the detection of autoantibodies. Of the 19 proteins measured by SRM, 12 were significantly decreased between IA and T1D post-IA (**Fig. 2B**) at a p-value threshold of 0.05 with a consistent pattern is also observed prior to IA with 5 of the 19 complement proteins significantly decreased (**Fig. 2A**). MBL2 unique to the lectin pathway is the only protein in the complement pathways with increased abundance across the entire time course. No proteins specific to the alternative pathway are significant. The time-course plots for each SRM peptide with the difference over time by individual subject and data point are given in Supplemental Fig. 1 and Supplemental Fig. 2, respectively, for the pre- and post-IA groups.

**Fig. 2:**
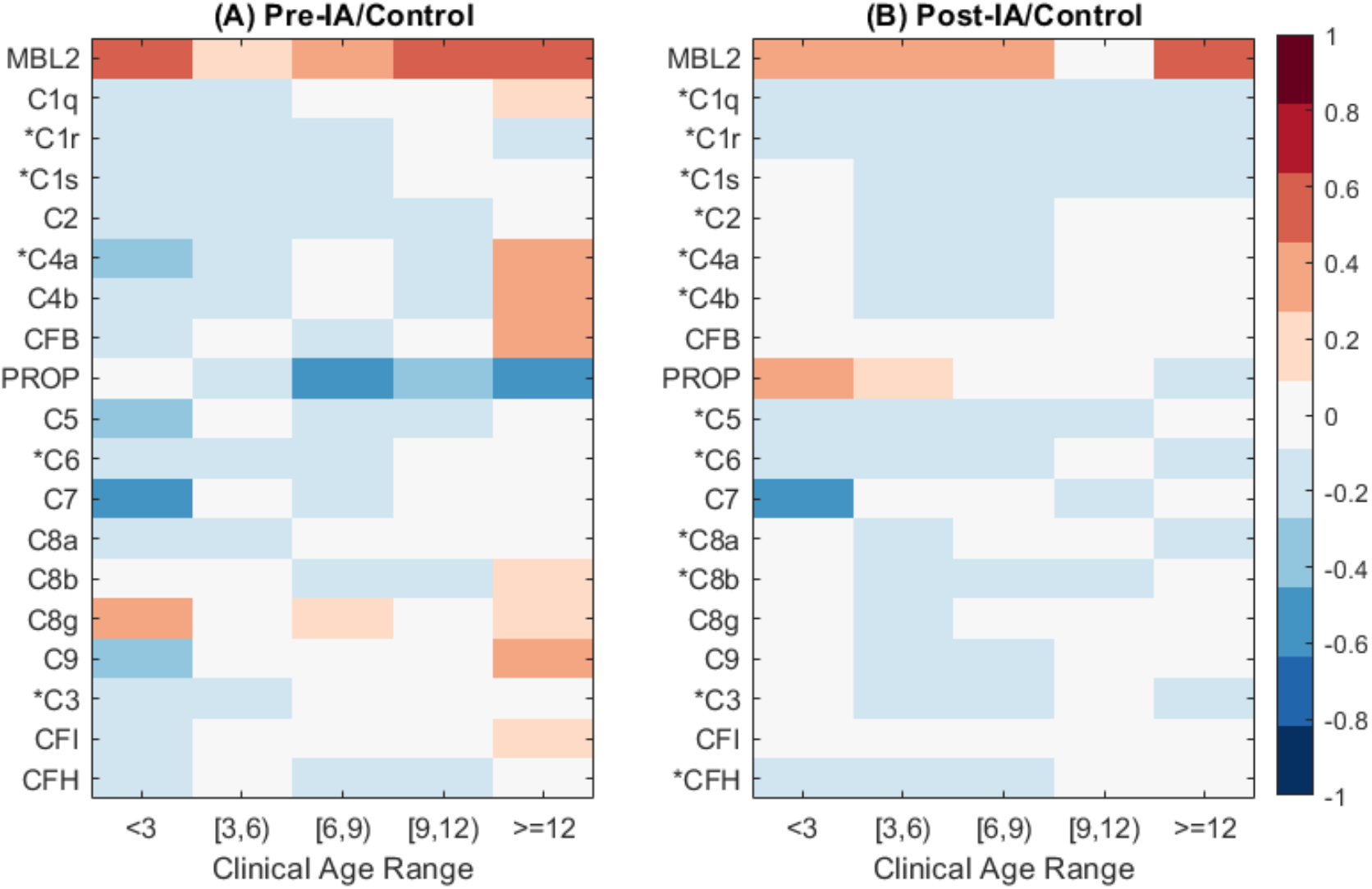
Complement Protein log2 fold-change for each statistical comparison of (A) pre-IA (n=47) and (B) post-IA (n=131). Proteins with an asterisk are significant at a *p-value* of 0.05 based on the full linear model.

To validate the SRM proteomics data in **Fig. 2**, we used previously collected data on 16 complement proteins using immunoassays (Exsera)^21^ of which 10 were in common with the SRM identified proteins. **Fig 3** shows the overall similarity in respect to log2 fold change between the two measurement types across all of the 10 common complement protein measurements made across all time points relative to control for both Pre- and Post-IA. C3 is the one complement protein that is significant (*p-value* < 0.05) for both the SRM and Exsera prior to IA and at or after IA. Of the remaining 9 proteins, prior to IA there is one (C4b) significant for SRM but not Exsera and one (MBL2) significant for Exsera but not SRM. For Post-IA, MBL2 is again significant for Exsera and although not statistically significant for SRM shows a similar pattern of increase relative to control. Similarly, C1q, C2, C4b, C5, CFH are significant for SRM, and although not statistically significant for Exsera show a common pattern of decreased abundance relative to control. Of the six proteins measured by immunoassays that do not overlap with the SRM dataset, three (C3a, C5a, C5b) were significantly decreased both for Pre-IA and Post-IA, again showing the same pattern of decreased abundance relative to controls. The presence of C3a and C5a is observed in both controls and cases, significant for the Exsera dataset^22–27^.

**Fig. 3:**
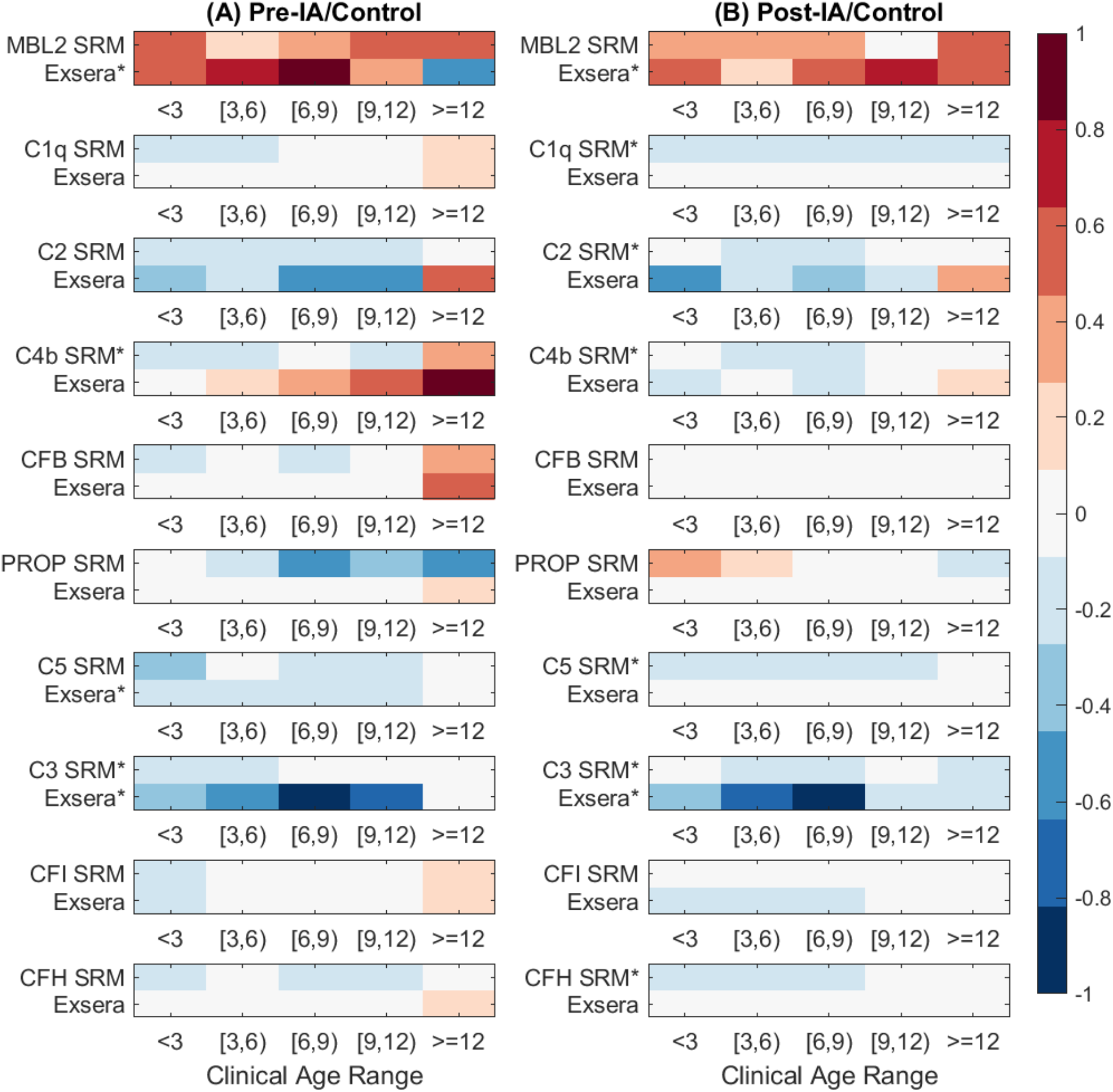
Average Exsera log2 fold-change in comparison to the average log2 fold change of the SRM measurements across time for the 10 proteins measured by both technologies. The * indicates that the protein measurement was significant based on the full linear model at a *p-value* of 0.05.

The effect size for each protein based on the statistical model measures the strength of the relationship between the protein abundance and the outcome. As seen in **Fig. 4**, most of the SRM and Exsera quantitatively measured proteins have a negative effect size, meaning that the effect of Pre-IA or Post-IA decreases on average relative to the controls. This matches with the observed log2 fold-changes in **Figs 2-3**. We observe that most proteins, with exception of MBL2, have a negative effect size. For the proteins measured by Exsera we observe that 68.8% and 87.5% of the proteins have a negative effect size for Pre- and Post-IA, respectively, similar to results observed for SRM. To evaluate the likelihood that this could be observed by chance, we simulated effect sizes from a uniform distribution randomly ranging from -1 to 1 for each of the 19 proteins and computed the proportion of negative values. We repeated this process 100 times and as expected the median was near 50%, specifically 47.4%. A Wilcoxon rank sum test was used to evaluate the null hypothesis that our random distribution could have the same median as our observed data. In all cases, our percentage of negative effect sizes was larger than expected by chance with a p-value less than 1.0E-7. We extended this analysis to also include a random p-value computational and evaluating the percentage with a p-value less than 0.05 and a negative effect. The likelihood of observing both the number of significant proteins and a negative directional change by chance is extremely small, less than 2.6E-20, both in the SRM and Exsera datasets.

**Fig. 4:**
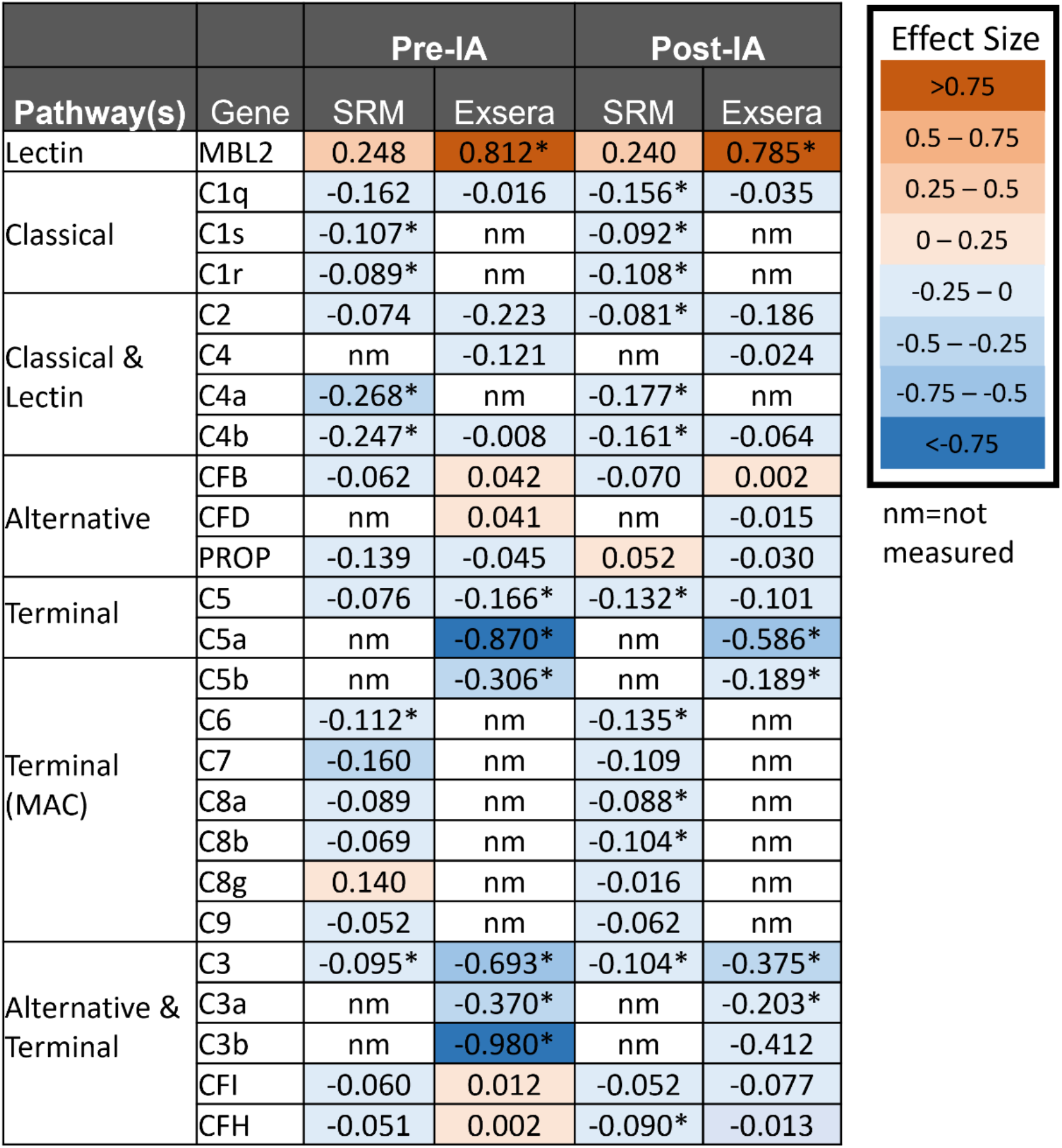
Effect size based on the statistical comparisons either prior to or at or after IA with proteins sorted based on complement pathway. Number in the boxes are the effect size from the linear mixed model comparing the difference of protein abundance between Control and the Pre-IA and Post-IA groups. A negative effect size indicates strength of the relationship between the protein abundance and the outcome compared, a positive effect means that Pre- or Post-IA increases the abundance on average relative to control and a negative effect is the contrary.

### Prediction of Progression based on Complement Proteins

To further explore the utility of the complement protein quantitation data to screen for children that will develop islet autoantibodies the 40 control and 47 Pre-IA children were down-selected to a single sample time point for machine learning. For the 40 control children this was the earliest sample collected and for the 47 Pre-IA children it was a random selection of the first or second time point prior to the detection of autoantibodies to assure the age distributions were not significantly different, average ages of the two groups are given in **Table 1**. Of the 47 children in the Pre-IA group 23 of them are diagnosed with T1D as of the last follow-up. All 25 proteins from both the SRM and Exsera measurements highlighted in **Fig. 4** were included in the initial dataset.

To identify the most predictive features a model-agnostic approach, Feature Importance Ranking Measure (FIRM)^28^ was utilized in conjunction with a linear Support Vector Machine (SVM). Analysis was performed using a 75/25 train/test split with repeated 10 fold cross-validation (CV)^29–31^. The accuracy of the model is quantified by a Receiver Operating Characteristic curve (ROC), specifically the area under this curve (AUC), for which the final average AUC as computed on the test data is 0.82. **Fig. 5A** gives the importance of each of the measured proteins from both the SRM and Exsera technologies ordered from the most important based on the results of FIRM. The most important features are C1r measured by SRM and C3a measured by Exsera, both of which are highly significant and as can be seen in **Fig 5B** visually separate the two groups fairly well. Evaluate of the data using an alternative approach, Recursive Feature Elimination performed repeatedly with 3-fold CV also found C1r was selected as the last feature to remove from the model in 100% of the RFE iterations and C3a was the second to the last to be removed in 99 of the 100 RFE iterations.

**Fig. 5:**
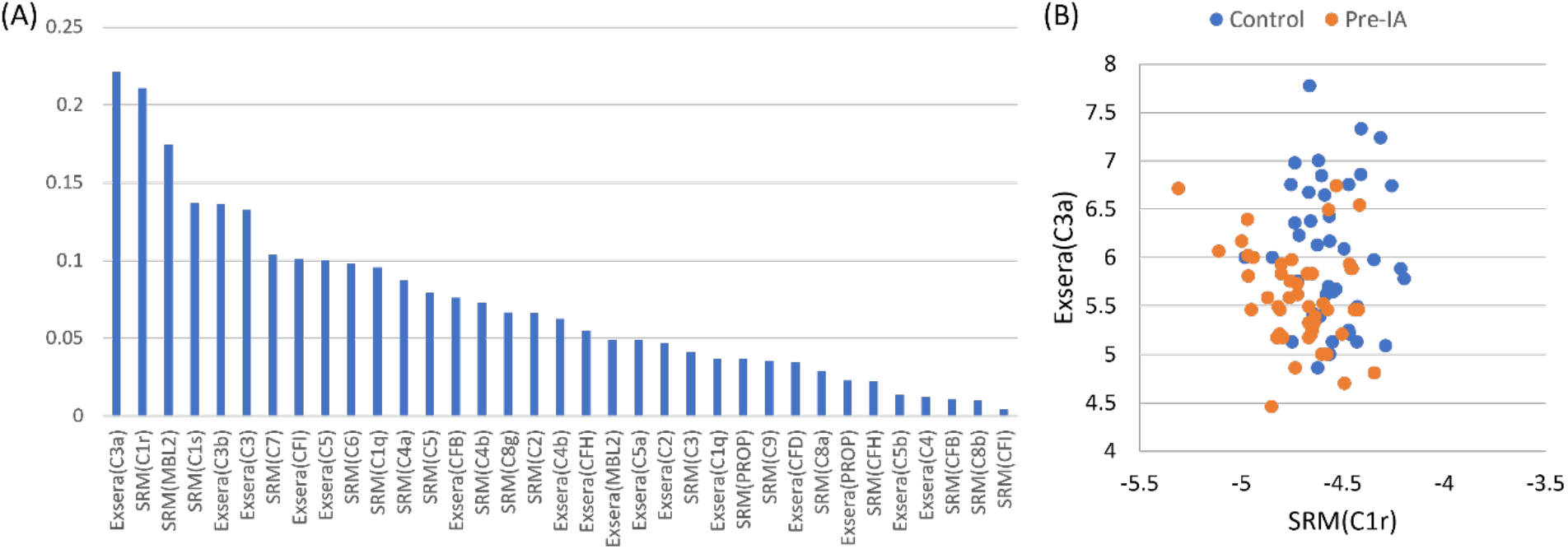
Machine learning results and scatter plot of the two most important features. (A) Feature importance values from most to least. (B) Scatter plot of the top two features shows a visual separation of control from Pre-IA.

## Discussion

There is an overwhelming pattern of a decrease in complement proteins prior to appearance of islet autoantibodies as well as after the seroconversion in children progressing to clinical diabetes in both the SRM and Exsera datasets. As seen in **Fig. 4**, except for MBL2, the overall pattern is a decrease across the entire set of complement proteins when compared to control. The classical and lectin pathways show the strongest decrease in complement proteins, observed both by statistical and machine learning analyses. This is consistent with prior findings of decreased abundance in specific complement proteins, such as C3 and C4, associated with T1D^32, 33^. The one distinct complement protein MBL2 is the lead of the lectin pathway and despite its increase, the C4a and C4b proteins are significantly decreased by SRM and show a similar decrease in Exsera, although not statistically significant. These findings for an increase in MBL2 related to onset of T1D have been observed previously^34, 35^. The membrane attack complex, also referred to as the terminal complement complex, is the result of the host’s complement system, and as seen in **Fig. 4**, all proteins in this complex are decreased, approximately half of them are significant at a p-value threshold of 0.05 for the three comparisons based on the results of the linear model. In general proteins associated with the Terminal pathway show a significant trend with 60% of the measurement significantly decreased. The Alternative pathway does not have any significant proteins and has mixed directional change based on the two types of measurements.

Complement levels are driven by several factors, including genetic polymorphisms driving transcriptional rates, responses to acute phase activators, and consumption through any of the three activation mechanisms. The observation that the activation products C5a, C3a and the membrane attack complex are all similarly decreased suggests that the causal relationships are likely not due to consumption by cleavage of the precursor molecules (C5 and C3) but rather to factors that regulate transcription, translation, and secretory pathways. Additional studies will be necessary to understand the causes of the decreases. An increase in complement activation has been shown to occur in pancreas of individuals with T1D^36^. However, it is unlike that deposition of complement proteins into islets would cause detectable reductions in levels of such abundant plasma proteins. Endogenous production of C3 by β cells is protective through an intracellular mechanism^37^. Exogenously added C3 protein also has a protective effect on human islets and rodent β cell line INS-1E against cytokine-induced apoptosis^14^. Therefore, it is likely that complement cascade represents protective mechanism for β cells, and that the lower levels of circulating complement proteins fail to provide protection against the autoimmune response. However, understanding how C3-mediated protection relates to systemic levels of complement proteins and whether lower circulating levels are reflective of an inadequate endogenous response to stress will require further study.

MBL2 is a pattern recognition molecule that engages the lectin pathway, and for which deficiencies are associated with infectious complications^38^. Why its levels would be discordant from the other complement factors is not readily understood but may reflect other functions of the molecule. In addition, the complement system as an innate immune pathway is intimately involved in the response to infection. In principle, a lower system could result in an inadequate response to pathogens or pathobionts that have been increasingly implicated in the development of T1D^39, 40^.

## Acknowledgements

This work was funded by NIH grant R01-DK32493 as well as The Leona M. and Harry B. Helmsley Charitable Trust grants 2018PG-T1D017 and G-2103-05121. Mass spectrometry analyses were performed in the Environmental Molecular Sciences Laboratory, a national scientific user facility sponsored by the Department of Energy (DOE) Office of Biological and Environmental Research and located at Pacific Northwest National Laboratory (PNNL). PNNL is operated by Battelle Memorial Institute for the DOE under contract DEAC05-76RLO1830. Identification of SRM targets was based on a large global LC-MS proteomics study supported by Environmental Determinants of Diabetes in the Young (TEDDY) consortium. The TEDDY Study is funded by U01 DK63829, U01 DK63861, U01 DK63821, U01 DK63865, U01 DK63863, U01 DK63836, U01 DK63790, UC4 DK63829, UC4 DK63861, UC4 DK63821, UC4 DK63865, UC4 DK63863, UC4 DK63836, UC4 DK95300, UC4 DK100238, UC4 DK106955, UC4 DK112243, UC4 DK117483, and Contract No. HHSN267200700014C from the National Institute of Diabetes and Digestive and Kidney Diseases (NIDDK), National Institute of Allergy and Infectious Diseases (NIAID), Eunice Kennedy Shriver National Institute of Child Health and Human Development (NICHD), National Institute of Environmental Health Sciences (NIEHS), Centers for Disease Control and Prevention (CDC), and JDRF. TEDDY is supported in part by the NIH/NCATS Clinical and Translational Science Awards to the University of Florida (UL1 TR000064) and the University of Colorado (UL1 TR002535).

## Author Contributions

B.M.W. performed statistical and machine learning analyses and wrote the manuscript. M.J.R led the study, interpreted results, and wrote the manuscript. E.S.N and T.O.M identified the SRM panel and interpreted results. A.S., Y.G., and T.F. performed SRM proteomics analyses. A.F. and V.M.H performed the Exsera proteomics analyses and interpreted results. F.D. and K.C.W designed the study and performed computational analyses. S.O. and S.S.R conceptualized the study and evaluated results. J.F and L.M.B generated the machine learning results and data resource. All authors revised and approved the manuscript.

## Methods

### DAISY Study cohort

DAISY follows prospectively 2,547 children at increased risk for T1D. The cohort consists of first-degree relatives of patients with type 1 diabetes and general population children with T1D susceptibility HLA *DR-DQ* genotypes identified by newborn screening^18^, recruited between 1993 and 2004. Follow-up results are available through April 4, 2022. Written informed consent was obtained from subjects and parents. The Colorado Multiple Institutional Review Board approved all protocols.

Autoantibodies were tested at 9, 15, and 24 months and, if negative, annually thereafter; autoantibody-positive children were retested every 3–6 months.

Radiobinding assays for insulin (IAA), GAD (GADA), insulinoma-associated protein 2 (IA-2A), and/or zinc transporter 8 (ZnT8A) autoantibodies were conducted as previously described^41, 42^. Subjects were considered persistently islet autoantibody positive if they had two or more consecutive confirmed positive samples, not due to maternal islet autoantibody transfer, or had one confirmed positive sample and developed diabetes prior to next sample collection. Diabetes was diagnosed using American Diabetes Association criteria.

### Targeted Proteomic Measurements

Protein digestion was carried out in an Eppendorf epMotion 5075 Liquid Handler. Five microliters of plasma from each donor were loaded into 96-well plates and 45 μL of 8M urea in 50 mM NH4HCO3 was added to each sample. Samples were reduced by adding 5 μL of 100 mM dithiothreitol and shaking at 1200 rpm for 1 h at 37 °C. Samples were alkylated by adding 5.5 μL of 400 mM iodoacetamide and shaking at 1200 rpm in the dark for 1 h at 37 °C. Samples were diluted by adding 300 μL 50 mM NH4HCO3 and were supplemented with 1 M CaCl2 to a final concentration of 1 mM and trypsin (Promega Sequencing Grade Modified Trypsin) to a final ratio of 1/50 (enzyme/protein). Proteins were digested for 6 h at 37 °C with shaking at 1200 rpm. Reactions were quenched by adding 10% trifluoroacetic acid to a final concentration of 0.1%. Samples were desalted in C18 solid phase extraction plates (Phenomenex) and dried in a vacuum centrifuge. Samples were dissolved in 100 μL of water and assayed with BCA (Thermo Fisher) to determine peptide concentration. Peptides were spiked with internal standards comprised of synthetic versions of the targeted peptides with heavy-isotope labeled amino acid residues at their C-termini (Vivitide, previously known as New England Peptide) and diluted to 0.2 μg/μL for mass spectrometry analysis. Assays were tested in different ratios of human and chicken plasma to ensure they were in linear response range.

Two microliters of peptides were loaded into a reverse phase column (Peptide BEH C18, 130 A 1.7um 0.1×100mm, Waters) connected to an Acquity M-Class Nano UHPLC system (Waters). The column temperature was set at 45 °C and peptides were separated with a gradient of water (solvent A) and acetonitrile (solvent B) both containing 0.1% formic acid. Eluting peptides were analyzed online by selected-reaction monitoring (SRM) in a triple quadrupole mass spectrometry (TSQ Altis, Thermo Fisher). The electrospray voltage was set to 2.1 kV and the source temperature at 350 °C. The LC-SRM raw data are available on MassIVE (https://massive.ucsd.edu); MSV000090848. Data quality was monitored using an in-lab developed tool name Q4SRM^43^.

All the LC-SRM data were imported into the Skyline software (MacLean et al., 2010) and the peak boundaries were manually inspected to ensure correct peak assignment and peak boundaries. There were 333 peptides representing 169 proteins measured in the final assay, and each peptide were monitored by 2-3 precursor-fragment ion pairs (i.e., transitions). The information about specific transitions were deposited within the Skyline files and can be accessed at https://panoramaweb.org/DAISY_SRM_PNNL.url. Peak detection and integration were determined based on two criteria: 1) the same LC retention time and 2) approximately the same relative peak intensity ratios across multiple transitions between the endogenous peptides and heavy isotope-labelled internal peptide standards. The total peak areas of endogenous peptides and their ratios to the total peak areas of the corresponding heavy isotope-labelled internal peptide standards were exported directly from the Skyline software. No further data manipulation was performed. This information can also be found with the deposited Skyline files.

### Exsera Complement Factor Measurements

Immunological complement analysis at Exsera BioLabs was performed in plasma that had not been previously thawed by a combination of multiplex and single assay methods. For the multiplex analysis the human complement bead-based xMAP technology (Luminex Corp, Northbrook IL) and commercially available kits (EMD Millipore, Milliplex Map, Burlington, MA) were used to measure thirteen complement proteins, spanning all three activation arms and the terminal pathway of complement. Measurements were made on a MagPix Luminex instrument. The Millipore Panel #1 was used to measure C2, C4b, C5a, C9, FD, MBL and Factor I (FI). Panel #2 was used to measure C1q, C3, C3b & iC3b, C4, FB, Factor H (FH), and P. In addition, the complement activation markers Bb, C3a and the soluble terminal complement complex, sC5b-9 were measured by ELISA (Quidel Corp, San Diego CA). All testing methods had been optimized and validated within Exsera BioLabs, a College of American Pathologists (CAP) and Clinical Laboratory Improvement Amendments (CLIA) certified laboratory

All analysis was performed in duplicate with the resulting mean values report. For the multiplex Luminex data the mean fluorescent intensity was the raw value and for the ELISA analysis the raw values were optical density. Standard curves were utilized with a four-parameter parametric curve fit used to calculate the absolute quality in ng/mL or mg/mL, as appropriate. Three quality controls (QC) were included in each run, including at least one laboratory developed and characterized QC. The QCs were monitored for performance and for all testing in the study the values returned were within required parameter, demonstrating assay performance. No further data manipulation was performed. Human reference ranges for the analytes tested have been determined within Exsera by the measurement of normal individuals.

### Statistical Analyses

A linear mixed model comparing the difference of peptide or protein abundance between CTRL and the Pre-IA and Post-IA groups, adjusting for sex, HLA group, and first-degree relative status with a nested random effect for subject and plate number was performed^44^. Statistical analysis was performed in MatLab® using the ‘fitglme’ function, from which p-values and effect size can be extracted as output arguments. The average log2 abundance values of all Pre-IA, Post-IA and control samples for each subject within the age range was used to compute log2 fold-changes. For all comparisons, the original data is available in Supplemental Dataset 1 and the final statistics, effect sizes and log2 fold-changes are given on the statistics tabs in the Supplemental Dataset 2.

### Machine Learning

A linear support vector machine (SVM) was employed for machine learning with model agnostic feature importance ranking metrics generated by Feature Importance Ranking Measure (FIRM)^28^ using the tidymodels R package. The choice of the hyperparameters for the SVM was determined as the combination that yielded the highest average Area Under a Receiver Operator Curve (AUC) across repeated 10-fold cross-validation. The final parameters used for the SVM was a margin of 09 and cost of 0.0132. To rank the features, FIRM is based on Individual Conditional Expectation (ICE) curves^45^. Briefly, ICE curves measure the effect of a predictor (or small subset of predictors) on the estimated prediction surface. Predictors that have no effect on the response correspond to flat ICE curves. Thus, to quantify importance, the FIRM approach averages the computed variances of the ICE values for a continuous predictor.

## Data Availability

The raw mass spectrometry data can be found at https://massive.ucsd.edu (MSV000090848) and the processed data is in Skyline (https://panoramaweb.org/DAISY_SRM_PNL.url). Source data used to generate all statistics and machine learning results are available upon reasonable request to the authors.

## Code Availability

For statistical analyses, all functions are available in Statistics and Machine Learning Toolbox in the MatLab platform. The machine learning analyses used functions available in R using the tidymodels package.

## References

1 Ziegler, A. G. et al. Seroconversion to multiple islet autoantibodies and risk of progression to diabetes in children. JAMA 309, 2473–2479, doi:10.1001/jama.2013.6285 (2013).

2 Kupila, A. et al. Feasibility of genetic and immunological prediction of type I diabetes in a population-based birth cohort. Diabetologia 44, 290–297, doi:10.1007/s001250051616 (2001).

3 Liu, X. et al. Distinct Growth Phases in Early Life Associated With the Risk of Type 1 Diabetes: The TEDDY Study. Diabetes Care 43, 556–562, doi:10.2337/dc19-1670 (2020).

4 Roll, U. et al. Perinatal autoimmunity in offspring of diabetic parents. The German Multicenter BABY-DIAB study: detection of humoral immune responses to islet antigens in early childhood. Diabetes 45, 967–973, doi:10.2337/diab.45.7.967 (1996).

5 Armento, A., Ueffing, M. & Clark, S. J. The complement system in age-related macular degeneration. Cell Mol Life Sci 78, 4487–4505, doi:10.1007/s00018-021-03796-9 (2021).

6 Mao, X. et al. Tumour extracellular vesicle-derived Complement Factor H promotes tumorigenesis and metastasis by inhibiting complement-dependent cytotoxicity of tumour cells. J Extracell Vesicles 10, e12031, doi:10.1002/jev2.12031 (2020).

7 Giang, J. et al. Complement Activation in Inflammatory Skin Diseases. Front Immunol 9, 639, doi:10.3389/fimmu.2018.00639 (2018).

8 Sjowall, C., Mandl, T., Skattum, L., Olsson, M. & Mohammad, A. J. Epidemiology of hypocomplementaemic urticarial vasculitis (anti-C1q vasculitis). Rheumatology (Oxford) 57, 1400–1407, doi:10.1093/rheumatology/key110 (2018).

9 Moulder, R. et al. Serum proteomes distinguish children developing type 1 diabetes in a cohort with HLA-conferred susceptibility. Diabetes 64, 2265–2278, doi:10.2337/db14-0983 (2015).

10 Ajjan, R. A. & Schroeder, V. Role of complement in diabetes. Molecular immunology 114, 270–277, doi:10.1016/j.molimm.2019.07.031 (2019).

11 von Toerne, C. et al. Peptide serum markers in islet autoantibody-positive children. Diabetologia 60, 287–295, doi:10.1007/s00125-016-4150-x (2017).

12 Liu, C. W. et al. Temporal expression profiling of plasma proteins reveals oxidative stress in early stages of Type 1 Diabetes progression. J Proteomics 172, 100–110, doi:10.1016/j.jprot.2017.10.004 (2018).

13 Torn, C. et al. Complement gene variants in relation to autoantibodies to beta cell specific antigens and type 1 diabetes in the TEDDY Study. Sci Rep 6, 27887, doi:10.1038/srep27887 (2016).

14 Dos Santos, R. S., et al. Protective Role of Complement C3 Against Cytokine Mediated β-Cell Apoptosis. Endocrinology 158, 2503–2521, doi:10.1210/en.2017-00104 (2017).

15 Atanes, P. et al. C3aR and C5aR1 act as key regulators of human and mouse beta-cell function. Cell Mol Life Sci 75, 715–726, doi:10.1007/s00018-017-2655-1 (2018).

16 Liu, C. W. et al. Temporal profiles of plasma proteome during childhood development. J Proteomics 152, 321–328, doi:10.1016/j.jprot.2016.11.016 (2017).

17 Zhang, Q. et al. Serum proteomics reveals systemic dysregulation of innate immunity in type 1 diabetes. J Exp Med 210, 191–203, doi:10.1084/jem.20111843 (2013).

18 Rewers, M. et al. Newborn screening for HLA markers associated with IDDM: diabetes autoimmunity study in the young (DAISY). Diabetologia 39, 807–812, doi:10.1007/s001250050514 (1996).

19 Frohnert, B. I. et al. Late-onset islet autoimmunity in childhood: the Diabetes Autoimmunity Study in the Young (DAISY). Diabetologia 60, 998–1006, doi:10.1007/s00125-017-4256-9 (2017).

20 Nakayasu, E. S. et al. Tutorial: best practices and considerations for mass spectrometry-based protein biomarker discovery and validation. Nat Protoc 16, 3737–3760, doi:10.1038/s41596-021-00566-6 (2021).

21 Prohaszka, Z., Nilsson, B., Frazer-Abel, A. & Kirschfink, M. Complement analysis 2016: Clinical indications, laboratory diagnostics and quality control. Immunobiology 221, 1247–1258, doi:10.1016/j.imbio.2016.06.008 (2016).

22 Burns, V. E., Edwards, K. M., Ring, C., Drayson, M. & Carroll, D. Complement cascade activation after an acute psychological stress task. Psychosom Med 70, 387–396, doi:10.1097/PSY.0b013e31816ded22 (2008).

23 Grumach, A. S., Ceccon, M. E., Rutz, R., Fertig, A. & Kirschfink, M. Complement profile in neonates of different gestational ages. Scand J Immunol 79, 276–281, doi:10.1111/sji.12154 (2014).

24 Lachmann, P. J. The amplification loop of the complement pathways. Adv Immunol 104, 115–149, doi:10.1016/S0065-2776(08)04004-2 (2009).

25 Norda, R. et al. Complement activation products in liquid stored plasma and C3a kinetics after transfusion of autologous plasma. Vox Sang 102, 125–133, doi:10.1111/j.1423-0410.2011.01522.x (2012).

26 Reis, E. S. et al. Sleep and circadian rhythm regulate circulating complement factors and immunoregulatory properties of C5a. Brain Behav Immun 25, 1416–1426, doi:10.1016/j.bbi.2011.04.011 (2011).

27 Yang, S., McGookey, M., Wang, Y., Cataland, S. R. & Wu, H. M. Effect of blood sampling, processing, and storage on the measurement of complement activation biomarkers. Am J Clin Pathol 143, 558–565, doi:10.1309/AJCPXPD7ZQXNTIAL (2015).

28 Scholbeck, C. A., Molnar, C., Heumann, C., Bischl, B. & Casalicchio, G. Sampling, Intervention, Prediction, Aggregation: A Generalized Framework for Model-Agnostic Interpretations. Comm Com Inf Sc 1167, 205–216, doi:10.1007/978-3-030-43823-4_18 (2020).

29 Frohnert, B. I. et al. Predictive Modeling of Type 1 Diabetes Stages Using Disparate Data Sources. Diabetes 69, 238–248, doi:10.2337/db18-1263 (2020).

30 Webb-Robertson, B. M. et al. Prediction of the development of islet autoantibodies through integration of environmental, genetic, and metabolic markers. J Diabetes 13, 143–153, doi:10.1111/1753-0407.13093 (2021).

31 Webb-Robertson, B. M. et al. Integration of Infant Metabolite, Genetic and Islet Autoimmunity Signatures to Predict Type 1 Diabetes by 6 Years of Age. J Clin Endocrinol Metab, doi:10.1210/clinem/dgac225 (2022).

32 Charlesworth, J. A. et al. The Complement-System in Type-1 (Insulin Dependent) Diabetes. Diabetologia 30, 372–379, doi:Doi 10.1007/Bf00292537 (1987).

33 Mason, M. J. et al. Low HERV-K(C4) Copy Number Is Associated With Type 1 Diabetes. Diabetes 63, 1789–1795, doi:10.2337/db13-1382 (2014).

34 Bouwman, L. H. et al. Elevated levels of mannose-binding lectin at clinical manifestation of type 1 diabetes in juveniles. Diabetes 54, 3002–3006, doi:DOI 10.2337/diabetes.54.10.3002 (2005).

35 Hansen, T. K. et al. Elevated levels of mannan-binding lectin in patients with type 1 diabetes. J Clin Endocrinol Metab 88, 4857–4861, doi:10.1210/jc.2003-030742 (2003).

36 Rowe, P. et al. Increased complement activation in human type 1 diabetes pancreata. Diabetes Care 36, 3815–3817, doi:10.2337/dc13-0203 (2013).

37 King, B. C. et al. Complement Component C3 Is Highly Expressed in Human Pancreatic Islets and Prevents beta Cell Death via ATG16L1 Interaction and Autophagy Regulation. Cell Metab 29, 202–210 e206, doi:10.1016/j.cmet.2018.09.009 (2019).

38 Garred, P., Larsen, F., Seyfarth, J., Fujita, R. & Madsen, H. O. Mannose-binding lectin and its genetic variants. Genes Immun 7, 85–94, doi:10.1038/sj.gene.6364283 (2006).

39 Vehik, K. et al. Prospective virome analyses in young children at increased genetic risk for type 1 diabetes. Nat Med 25, 1865–1872, doi:10.1038/s41591-019-0667-0 (2019).

40 Oikarinen, S. et al. Characterisation of enterovirus RNA detected in the pancreas and other specimens of live patients with newly diagnosed type 1 diabetes in the DiViD study. Diabetologia 64, 2491–2501, doi:10.1007/s00125-021-05525-0 (2021).

41 Gianani, R. et al. ICA512 autoantibody radioassay. Diabetes 44, 1340–1344 (1995).

42 Wenzlau, J. M. et al. The cation efflux transporter ZnT8 (Slc30A8) is a major autoantigen in human type 1 diabetes. Proc Natl Acad Sci U S A 104, 17040–17045, doi:10.1073/pnas.0705894104 (2007).

43 Gibbons, B. C. et al. Rapidly Assessing the Quality of Targeted Proteomics Experiments through Monitoring Stable-Isotope Labeled Standards. J Proteome Res 18, 694–699, doi:10.1021/acs.jproteome.8b00688 (2019).

44 McCulloch, C. E., Searle, S. R. & Neuhaus, J. M. *Generalized, Linear, and Mixed Models*. 2nd edn, (Wiley-Interscience, 2008).

45 Goldstein, A., Kapelner, A., Bleich, J. & Pitkin, E. Peeking Inside the Black Box: Visualizing Statistical Learning With Plots of Individual Conditional Expectation. J Comput Graph Stat 24, 44–65, doi:10.1080/10618600.2014.907095 (2015).

